# A generic, scalable, and rapid TR-FRET –based assay for SARS-CoV-2 antigen detection

**DOI:** 10.1101/2020.12.07.20245167

**Authors:** Juuso Rusanen, Lauri Kareinen, Leonora Szirovicza, Hasan Uğurlu, Lev Levanov, Anu Jääskeläinen, Maarit Ahava, Satu Kurkela, Kalle Saksela, Klaus Hedman, Olli Vapalahti, Jussi Hepojoki

## Abstract

The ongoing COVID-19 pandemic has seen an unprecedented increase in the demand for rapid and reliable diagnostic tools, leaving many laboratories scrambling for resources. We present a fast and simple method for the detection of SARS-CoV-2 in nasopharyngeal swabs. The method is based on the detection of SARS-CoV-2 nucleoprotein (NP) and S protein (SP) via time-resolved Förster resonance energy transfer (TR-FRET) with donor- and acceptor-labeled polyclonal anti-NP and -SP antibodies. Using recombinant proteins and cell culture-grown SARS-CoV-2 the limits of detection were established as 25 pg of NP or 20 infectious viral units (i.u.), and 875 pg of SP or 625 i.u. of SARS-CoV-2. Testing RT-PCR positive (n=48, with cycle threshold [Ct] values from 11 to 30) or negative (n=96) nasopharyngeal swabs, we showed that the assay yields positive results for all samples with Ct values of <25 and a single RT-PCR negative sample. We determined the presence of infectious virus in the RT-PCR-positive nasopharyngeal swabs by virus isolation, and observed a strong association between the presence of infectious virus and a positive antigen test result. The NP-based assay showed 97.4% (37/38) sensitivity and 100% (10/10) specificity in comparison with virus isolation, and 77.1% (37/48) and 99.0% (95/96) in comparison with SARS-CoV-2 RT-PCR. The assay is performed in a buffer that neutralizes SARS-CoV-2 infectivity and is relatively simple to set up as an “in-house” test. The assay principle as such is applicable to other viral infections, and could also be readily adapted to a massively high throughput testing format.

## INTRODUCTION

The ongoing coronavirus disease 2019 (COVID-19) pandemic has by December 2020 claimed almost one and a half million lives globally, with over 60 million confirmed infections. To manage the disease, accurate diagnostic tools are of key importance. Detection of the causative agent, the severe acute respiratory syndrome coronavirus 2 (SARS-CoV-2), or its parts is the cornerstone of diagnosis, as the disease presentation is often indistinguishable from other respiratory infections. The mainstay of COVID-19 diagnosis is RT-PCR testing, done typically from a nasopharyngeal swab (NPS), while also oropharyngeal or mid-turbinate swab as well as salivary samples are in use. Alternatively, the less labour-intensive antigen detection tests are also increasingly deployed. Antigen tests tend to be specific, but analytically less sensitive than RT-PCR. RT-PCR can detect viral nucleic acid even after the infectious virus has waned, with the individual at this time unlikely posing a transmission risk^1-3^. Evidence suggests that antigen testing may correlate with recovery of infectious virus better than a binary RT-PCR^4^. Frequent antigen testing has been proposed as an alternative approach in reducing community transmission of SARS-CoV-2^5^.

SARS-CoV-2 is an enveloped (+)ssRNA virus of genus *Betacoronavirus* in family *Coronaviridae* of the *Nidovirales* order. It contains four structural proteins: the nucleoprotein (NP) forms a ribonucleoprotein complex with the 30 kb non-segmented viral genome. The envelope (E) and membrane (M) proteins are embedded in the envelope, as is the spike protein (SP), protruding from the virion surface and generating large surface projections termed the corona. The SP undergoes processing to yield S1, which contains the receptor-binding domain (RBD) initially attaching the virus to angiotensin-converting enzyme-2 (ACE-2) on the host cell membrane, and S2 that mediates virus-cell fusion. In response to the pandemic, dozens of commercial SARS-CoV-2 antigen tests are available, predominantly of lateral flow or enzyme immunoassay type. Most target NP as analyte^6^. Of the seven antigen tests having by December 2020 received emergency use authorization (EUA) from the U.S. Food and Drug Administration (FDA), six target N and one S^7^.

Over the past few years, we have actively employed time-resolved Förster resonance energy transfer (TR-FRET) as basis of rapid homogeneous “mix and read” immunoassays for antibody detection^8-14^. FRET occurs when a donor and an acceptor fluorophore are in close proximity, whereby the excited donor transfers energy to the acceptor, which then emits a photon at a distinct wavelength. The closer the donor and acceptor are, the more frequent is the energy transfer, with a 50% efficiency typically achieved at a distance of 15 to 60 Å. Chelated lanthanide donor-enabled TR-FRET allows for measurement from autofluorescent biological samples.

Herein, we describe a rapid TR-FRET -based method for detection of SARS-CoV-2 NP and SP. In the assay polyclonal anti-NP and anti-RBD rabbit antibodies, each labeled with either a donor or an acceptor fluorophore, are combined at equimolar ratio and mixed with the clinical sample. The antigen, if present, binds the labeled antibodies and brings the fluorophores to close proximity. This results in a TR-FRET signal upon excitation, indicating presence of the antigen. We initially demonstrate the limits of detection for recombinant NP and SP as well as cell culture-grown SARS-CoV-2. We then evaluate the assay performance among 48 RT-PCR positive and 96 negative clinical NPS samples, and compare the antigen detection results to those of RT-PCR and virus cultivation.

## MATERIALS AND METHODS

### Patient samples and reference results

The evaluation of the SARS-CoV-2 TR-FRET assay was conducted by using nasopharyngeal swab (NPS) specimens collected in saline. The specimens were retrieved from patients with a clinically suspected COVID-19 and they were originally sent to HUS Diagnostic Center, HUSLAB for SARS-CoV-2 RT-PCR testing. The specimens were subsequently stored at −20°C.

The SARS-CoV-2 RT-PCR was based on a laboratory-developed test (LDT). The details and performance of the test in our laboratory setting have been described previously^15^. In this method (based on the N-gene, modified from Corman et al.^16^), the specimens were inactivated by combining 250 μl of MagNA Pure Lysis/Binding Buffer (Roche Diagnostics GmbH, Mannheim, Germany) and 250 μl of the specimen. Nucleic acid extraction was done from 450 μl specimen lysate with the MagNA Pure Viral NA SV 2.0 Kit (Roche Diagnostics GmbH, Mannheim, Germany). RT-PCR was performed using the SuperScript III Platinum One-Step qRT-PCR Kit with 600 nM of the forward primer CACATTGGCACCCGCAATC, 800 nM of the reverse primer GAGGAACGAGAAGAGGCTTG and 200 nM of the probe FAM-ACTTCCTCAAGGAACAACATTGCCA-BBQ3.

The SARS-CoV-2 RT-PCR positive panel comprised 48 specimens with cycle threshold (Ct) values ranging linearly between 11.42 and 29.98 in the LDT. The SARS-CoV-2 RT-PCR negative panel comprised 96 samples negative in the LDT. Patient data were collected and samples handled according to research permit approved by the local review board, permit HUS/32/2018 (Helsinki University Hospital, Finland).

### Cell lines, virus isolation and propagation

Vero E6 cells were transduced with a lentiviral vector expressing human Transmembrane serine protease 2, TMPRSS2, transcript variant 2 cDNA (NM_005656.4) and as selection marker blasticidin. Specifically, 1 ml of 0.22 µm filtered (Millipore) infectious supernatant of HEK293T cells transfected on a 10-cm plate 48 h earlier using 30 µg polyethylenimine with 5 µg pLenti6.3/V5-DEST TMPRSS2 (obtained from Biomedicum Functional Genomics Unit, University of Helsinki), and 5 µg p8.9NDSB^17^ plus 2-5 µg of pMD2.G (a gift from Didier Trono, Addgene plasmid #12259; https://n2t.net/addgene:12259; RRID:Addgene_12259) was added on Vero E6 cells seeded onto 6-well plates. Following 2 days of selection with 15 µg/ml of Blasticidin S HCl (Invitrogen), the cells were allowed to expand until confluency, and were subcultured three times. Once confirmed p24 negative, a clonal population of Vero E6-TMPRSS2 cells was obtained by limiting dilution. The obtained clones (N=5) were analyzed for TMPRSS2 expression by immunoblotting with V5 antibody (Invitrogen). The clone expressing the highest amount of TMPRSS2, VE6-TMPRSS2-H10, was selected for use.

SARS-CoV-2 isolation from clinical samples (stored at −20°C since the day of collection, not subjected to freeze-thaw) was attempted on both Vero E6 and VE6-TMPRSS2-H10 cells. Both cell lines were cultivated in Minimal Essential Medium Eagle (MEM, Sigma) supplemented with 10% fetal bovine serum (FBS, Gibco), 100 IU penicillin and 100 µg/ml streptomycin (Sigma), and 2 mM L-glutamine (Sigma). For isolation, the cells were grown on 12-well plates until approximately 90% confluent, the growth medium replaced with 400 µl of MEM-2% (as above but with 2% FBS), followed by addition of 50 µl of the NPS sample (under biosafety level 3 conditions), and 1 h incubation at 37°C 5% CO_2_. After two washes with MEM-2%, cultures were kept in 1 ml fresh MEM-2% for 4 days at 37°C (5% CO_2_), the medium collected and clarified by centrifugation (3,000 x rcf, 5 min), and the cells fixed for 15 min at room temperature by 3.7% formaldehyde in phosphate-buffered saline (PBS) followed by PBS wash and UV inactivation (5,000 x100 mJ, UV Crosslinker, CL-1000, Jena Analytik). The fixed cells were Crystal violet stained, and the extent of cytopathic effect (CPE) scored from 0 to 3 (from non-observable to extensive cell death). To confirm infection, RNA was extracted (QIAgen QIAamp Viral RNA Extraction Kit, following manufacturer’s protocol) from 100 µl of each cell culture supernatant, and the presence or absence of SARS-CoV-2 analyzed by RT-PCR targeting the RdRp (RNA-dependent RNA polymerase) gene as described^16^.

For TR-FRET antigen detection experiments, we produced a stock of SARS-CoV-2 in Vero E6 cells^18^. Briefly, 90-95% confluent Vero E6 cells were inoculated with 500 µl of 1:100 diluted SARS-CoV-2 containing supernatant (passage 7, approximately 5 × 10^7^ tissue culture infectious dose 50, TCID50, per ml). After 1 h of virus adsorption, the medium was replaced with MEM-2%, and after 2 days at 37°C 5% CO2, the supernatant collected and clarified by centrifugation (3,000 x rcf, 5 min), and stored in aliquots at −80°C. UV inactivation of culture supernatants was done as described above.

Human coronavirus 229E (hCoV-229E, kindly provided by Dr. Sisko Tauriainen, University of Turku, Turku, Finland) and NL63 (hCoV-NL63, kindly provided by Lia van der Hoek, Academic Medical Center, Amsterdam, Netherlands) served as controls for estimating cross-reactivity of the assay. The hCoV-229E stock was produced by inoculating rhesus macaque kidney cells, LLC-MK2 (from ATCC), with 500 µl of 1:1000 diluted cell culture supernatant (approximately 5 × 10^9^ TCID50/ml) 1 h at 37°C 5% CO2. After virus adsorption, the medium was changed into MEM-2%, the cells grown for 5 d (37°C, 5% CO2), the supernatant collected, centrifuged (3,000 x rcf, 5 min), and stored in aliquots at −80°C. The hCoV-NL63 stock was produced by inoculating human lung fibroblasts (MRC-5, from ATCC) with 500 µl of 1:100 diluted cell culture supernatants (approximately 1 × 10^6^ TCID50/ml) for 1 h at 37°C 5% CO2. After virus adsorption, the media were replaced with MEM-2%, the cells grown for 7 d (until appearance of definitive CPE), supernatants collected, centrifuged (3,000 x rcf, 5 min) and stored in aliquots at −80°C. The hCoV-229E and hCoV-NL63 supernatants were inactivated for the experiments by mixing at 1:10 in RIPA buffer (50 mM Tris-HCl pH 8.0, 150 mM NaCl, 1% NP-40, 0.1% SDS, 0.5% sodium deoxycholate and Roche cOmplete EDTA-free protease inhibitor cocktail).

### Antigens and antibodies

The production and purification of SARS-CoV-2 NP and SP antigens, followed a described protocol^14,19,20^. The RBD of the SP was produced in Expi293F cells as described^19,20^. Rabbit antisera against RBD and NP were generated at BioGenes GmbH (Berlin, Germany): day 0 initial dose 150 µg, day 7 booster 75 µg, day 14 booster 75 µg, day 28 booster 150 µg, and day 42 final bleed. For affinity purification, RBD and NP were coupled to CNBr-Sepharose 4B (Cytiva) following the manufacturer’s protocol. The respective antisera were passed through coupled Sepharoses packed into Poly-Prep Chromatography Columns (Bio-Rad), washed with 20 column volumes of phosphate-buffered saline (PBS), eluted (0.1 M glycine, 150 mM NaCl, pH 2.5) with 2 M Tris, pH 9.0, concentrated using Amicon Ultra 15 ml 100 kDa-NMWL Centrifugal Filter (Millipore/Merck), and dialyzed against PBS using Slide-A-Lyzer Dialysis Cassettes (Thermo Scientific).

### Labelling

We labelled the affinity-purified antibodies, 250 µg/reaction, with donor (europium, Eu) and acceptor (Alexa Fluor 647, AF647) respectively using QuickAllAssay Eu-chelated protein labeling kit (BN Products and Services Oy) and Alexa Fluor™ 647 NHS Ester (Thermo Scientific) following the manufacturer’s instructions. Disposable PD-10 Desalting Column with Sephadex G-25 resin (Cytiva) served to remove non-reacted fluorophores, and Amicon Ultra 0.5 ml 50 kDa-NMWL Centrifugal Filter (Millipore/Merck) for concentrating the labelled antibodies, which were then stored aliquoted at −80°C until use.

### TR-FRET assays

First, we set up TR-FRET assays for SARS-CoV-2 SP and NP antigens, by using the respective purified proteins as well as the corresponding Eu- and AF-labeled anti-RBD and anti-NP antibodies. The assay principle and workflow are depicted in Fig. 1. Briefly, antibody mixes with equimolar concentrations of Eu- and AF-labeled anti-RBD and anti-NP antibody concentrations were prepared in RIPA buffer. For setting up the assay, a pool of four SARS-CoV-2 negative NPS samples was divided in aliquots and spiked with either NP or SP proteins at various concentrations. 10 µl of antibody mix was pipetted on a 384-well microplate (ProxiPlate 384 Plus F, Black 384-shallow well microplate, PerkinElmer, USA), followed by 10 µl of the antigen-spiked sample. TR-FRET signal was measured directly thereafter and at time points 7, 15, 22, 30, 45, 60 and 90 minutes after the first measurement with a Hidex Sense microplate reader (Hidex Oy, Finland). FRET donor excitation was done at 330 nm and after a delay of 70 µs the donor and acceptor signals were measured for 100 µs at 616 and 665 nm, respectively. TR-FRET signals were expressed as HTRF ratios, calculated as follows: HTRF ratio = emission at 616 nm / emission at 665 nm x 10000. Thereafter, the HTRF ratios measured from the antigen-spiked samples were compared with those measured from a non-spiked sample in the same run to calculate the fold increase in HTRF ratio. Antibody plate concentrations ranging from 5 to 500 nM (half Eu- and half AF-labeled) were cross-titrated with antigen plate concentrations ranging from 5 nM to 500 nM.

**Figure 1.**
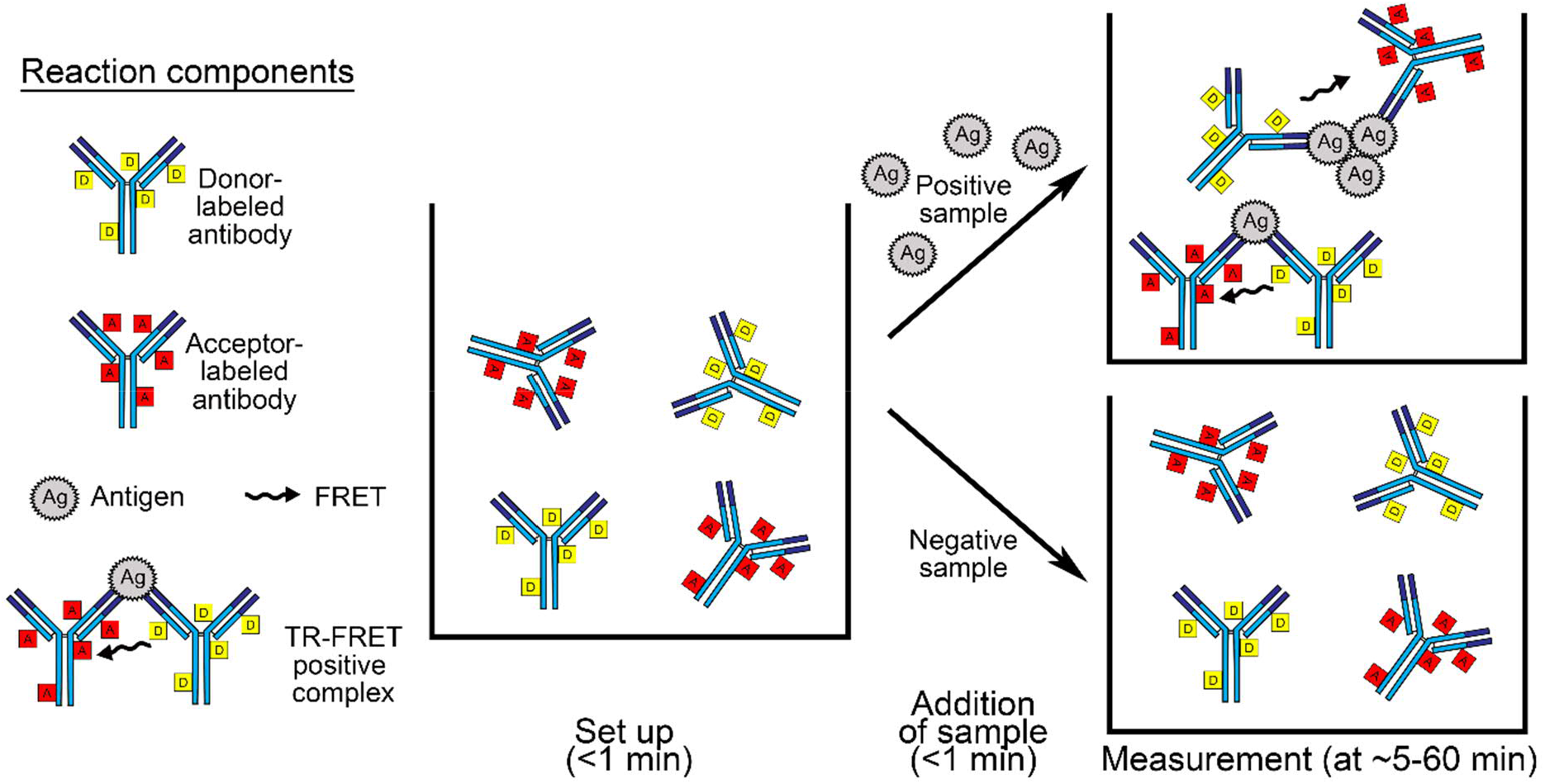
The TR-FRET assay workflow and principle. The left side shows the reaction components. At the center is a well containing donor- and acceptor-labeled antibodies at 1:1 molar ratio in the reaction buffer, in our set up the total antibody concentration at this point is 24 nM and the volume 10 µl. The arrows indicate addition of the sample material, in our set up either 10 µl of purified recombinant protein or 10 µl of NPS sample. The top right side shows schematically the antigen-antibody complexes formed following addition of sample containing the antigen, reaction volume at this stage in our set up is 10 µl. The bottom right side schematically demonstrates that the labelled antibodies do not form TR-FRET active complexes in absence of the antigen.

The ranges of antigen concentrations detectable by TR-FRET (at Eu- and AF-labeled anti-NP/-RBD concentrations of 6 + 6 nM) were then assessed by performing the assay as described above using N and S plate concentrations of 5 fM to 5 nM.

To assess the assay performance with samples containing virions, cell culture supernatants containing roughly 10^7^ TCID50/ml of SARS-CoV-2 were used. For initial experiments (carried out in BSL-3 laboratory) infectious cell culture supernatant (undiluted, 1:10, 1:25, 1:50, and 1:100 diluted in RIPA) was used. After verifying that UV inactivated virus produced similar results, the negative NPS sample matrix was spiked with UV-inactivated virus-containing cell culture supernatant to yield a dilution series from 1:10 to 1:20480. The samples were tested in the TR-FRET assays performed as above at Eu- and AF-labeled anti-NP/-RBD concentrations of 6 + 6 nM.

The NPS sample analysis was done by mixing 10 µl of the sample with 10 µl of the antibody mixes (Eu- and AF-labeled anti-NP/-RBD concentrations of 6 + 6 nM). The TR-FRET assays with SARS-CoV-2 RT-PCR positive NPS samples were carried out in BSL-3 laboratory and RT-PCR negative samples in BSL-2 laboratory. The signals produced by hCoV-229E and hCoV-NL63 were evaluated by mixing 10 µl (undiluted, 1:10, 1:25, 1:50, and 1:100 diluted in RIPA) of the cell culture supernatant with 10 µl of the antibody mixes (at Eu- and AF-labeled anti-NP/-RBD concentrations of 6 + 6 nM).

## RESULTS

### Proof-of-concept for the homogeneous antigen detection assay

We hypothesized that homogenous i.e. in solution detection of antigens could be achieved utilizing a polyclonal antibody separately labelled with fluorophores forming a FRET pair. To test the hypothesis and assay principle presented in Fig. 1, we generated antisera against SARS-CoV-2 NP and the RBD of SP, titers >204,800 based on NP and SP ELISA respectively. Following affinity purification, the antibodies were labelled with chelated europium (Eu, donor) and AlexaFluor 647 (AF647, acceptor). In the first experiments, we tested the assay principle by mixing recombinant NP and SP with 1 µM (500 nM nM Eu labelled + 500 nM AF647 labelled) anti-NP and anti-RBD antibody mixtures in the presence of increasing bovine serum albumin (BSA) concentrations. Addition of BSA increased the signal-to-noise ratio, encouraging us to evaluate the assay performances further utilizing infectious SARS-CoV-2 containing cell culture supernatants and different buffer compositions. The assay with 1 µM antibody concentrations produced respectable signal-to-noise ratios in detergent-containing RIPA (radioimmunoprecipitation assay) buffer (Fig. S1). RIPA buffer (refer to materials and methods for the full recipe) used here contained 1% NP-40 and 0.1% SDS, both shown to effectively inactivate the enveloped SARS-CoV-2^21,22^, which is why we to employ RIPA for the subsequent analyses.

### Assay optimization using recombinant antigens and SARS-CoV-2

To optimize the assay performance, we mixed the labelled antibodies at equimolar ratio with known amounts of recombinant NP and SP, and recorded the produced TR-FRET signals (as HTRF, homogeneous time-resolved fluorescence, values) as a function of time. The results showed the assays to produce the highest HTRF values when the concentration of labeled antibodies equalled the concentrations of the purified respective antigens (Fig. S2). Higher antibody concentrations shortened the time required for reaching the signal peak (fold increase in HTRF value, HTRF_sample_/HTRF_buffer_): In an antibody concentration of 5 nM (2.5 nM Eu-+ 2.5 nM of AF647-labeled) it took ∼60 min for the signal to peak for both NP and SP, whereas in an antibody concentration of 500 nM, the NP signal peak occurred in 7 min and the SP signal peaked in ∼30 minutes. We also tested the assay performances with Eu- and AF-labeled antibodies mixed at the unequal proportions of 1:2 and 2:1, but this did not increase the signal to background ratio (Table S1).

We then evaluated the assay performance at 50, 25, 12, and 6 nM total antibody concentrations using SARS-CoV-2 containing cell culture supernatants at different dilutions. We included a UV-inactivated cell culture supernatant to find out whether UV inactivation would affect the analysis. The results concurred with the findings using recombinant antigens, and showed the dependence of the TR-FRET signal kinetics on the antigen concentration (Fig. S3). The results further indicated that 12 nM total antibody concentration enables measurement of antigens over a broad range of virus concentrations, and that infectious and UV-inactivated SARS-CoV-2 produce similar results.

### Limits of detection

To assess the limits of detection (LOD) for the assays at 12 nM total antibody concentration, we spiked a pool of NPS samples with either purified antigens or inactivated SARS-CoV-2 at different dilutions. With purified antigens, the lowest concentrations producing readily detectable signals were 0.05 nM for NP and 0.5 nM for SP (Fig. S4, a-b). With UV-inactivated SARS-CoV-2, dilutions of the supernatant up to 1:5120 and 1:160 produced reliably measurable signals with anti-NP and anti-RBD antibodies, respectively (Fig. S4, c-d). With the 10 µl sample volume, the detection limit of the assay for recombinant NP was ∼25 pg (using molecular weight of 50 kDa) and 875 pg for the SP (using molecular weight of 175 kDa). Correspondingly, the NP assay could detect approximately 15 and the RBD assay approximately 420 plaque-forming units (converted by using 0.7 x TCID50/ml=pfu/ml) per reaction.

### Detection of SARS-CoV-2 antigens in NPS samples

After setting up the assay conditions using recombinant proteins and cell culture grown virus, we tested the assays in detection of viral antigen in SARS-CoV-2 RT-PCR positive NPS samples. We had 48 NPS samples with RT-PCR cycle threshold (Ct) values linearly ranging from ∼12 to 30 (Fig. S5), and employed total antibody concentrations of 50, 25, 12, and 6 nM. We observed that samples with Ct values of ≤25 yielded a signal in the NP assay (Fig. S6). By using the optimized conditions with 12 nM total labeled antibodies, the sensitivity of NP TR-FRET assay in comparison with RT-PCR was 77.1% (37/48). The SP assay showed greater diversity; most samples with Ct values ≤15 yielded a positive result (Fig. S6). We also performed an immunoblot to detect NP and SP in NPS samples covering the Ct value range of 12.8 to 26.2, and could detect SP and NP in samples with Ct value <22 (Fig. S7).

### Association between infectious virus and antigen detection

To determine to what extent the antigen assays correspond to the amounts of infectious virus in the sample, we subjected the 48 SARS-CoV-2 RT-PCR positive NPS samples to virus isolation. Transmembrane serine protease 2 (TMPRSS2) has been reported to function in priming the SARS-CoV-2 spike for entry^23^, and we thus chose to use both wild-type and a clonal population of TMPRSS2 expressing (VE6-TMPRSS2-H10) Vero E6 cells (Fig. S8). As indicated by cytopathic effects as well as RT-PCR from the cell culture supernatants, SARS-CoV-2 was isolated from 35/48 and 38/48 NPS samples with Vero E6 and VE6-TMPRSS2-H10 cells, respectively. Interestingly, the cell culture supernatants from VE6-TMPRSS2-H10 cells yielded a positive result 3 to 5 cycles earlier than did the supernatants from Vero E6 cells, pointing to ∼10-40 times more efficient virus production (Fig. S9). Altogether, infectious SARS-CoV-2 was recovered from all samples showing Ct values ≤24.5. We then compared the antigen assay to virus isolation from the respective samples, and observed all of the samples with Ct values ≤24.5 to be positive in the NP assay (Fig. 2a). Of the samples with Ct values >24.5 all produced a negative result in the TR-FRET NP assay, and SARS-CoV-2 was recovered from only one (Ct 24.87). By using the optimized conditions with 12 nM total labeled antibodies, the sensitivity of NP assay in comparison with virus isolation was 97.4% (37/38). The performance of the SP assay was poorer; only 8/38 samples with recoverable SARS-CoV-2 yielded a positive result (Fig. 2b).

**Figure 2.**
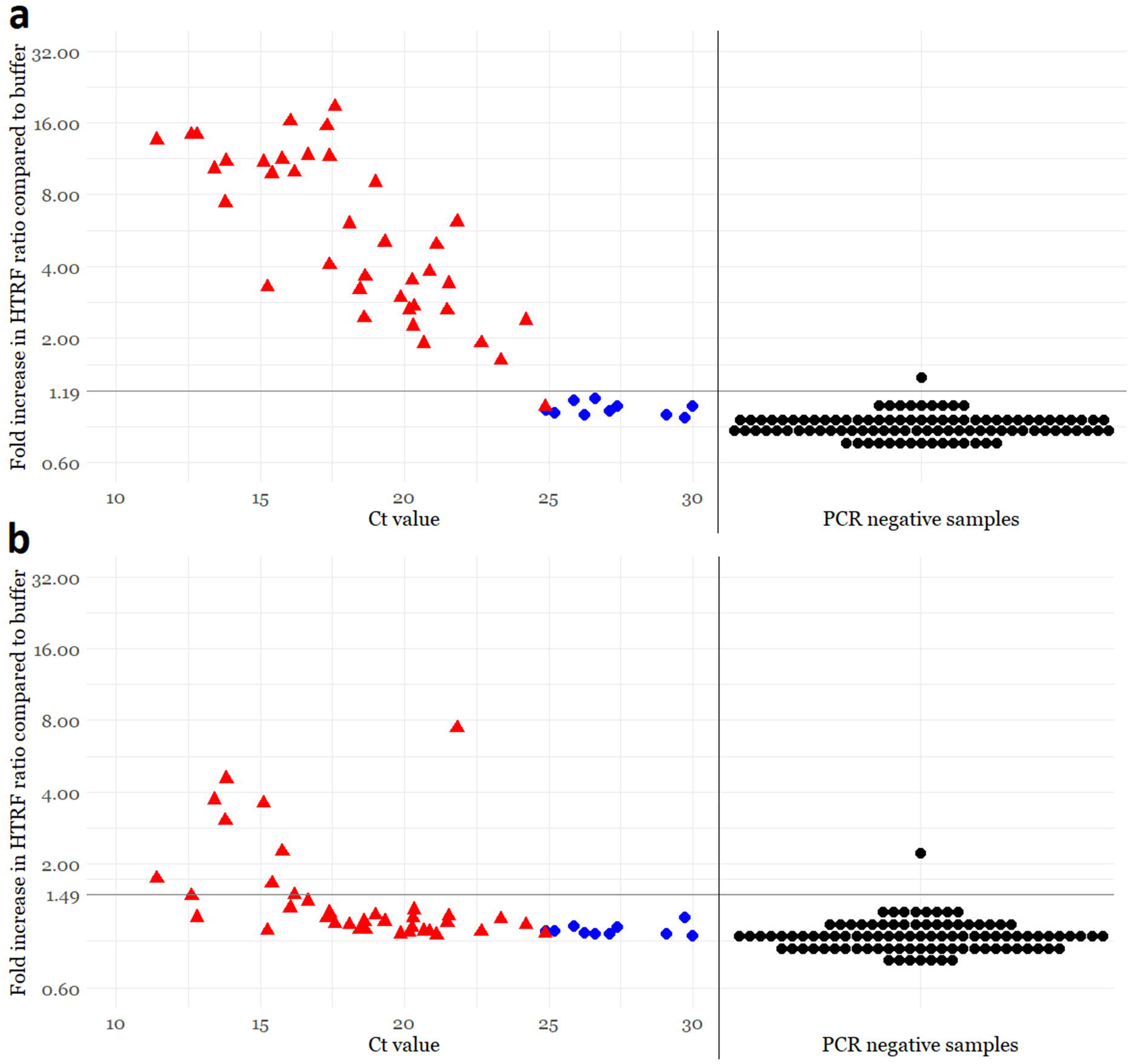
Comparison of TR-FRET based antigen detection and the amount of virus as analyzed by SARS-CoV-2 RT-PCR and virus isolation from NPS samples. The total antibody concentration in the assays is 12 nM (6 nM of Eu- and 6 nM of AF647-labeled antibody). a) Anti-NP assay results. b) Anti-RBD assay results. The y-axis (log scale) indicates a fold increase in HTRF ratio (HTRF_sample_/HTRF_buffer_) measured directly after pipetting the samples on plate. The x-axis shows the Ct value measured in the diagnostic SARS-CoV-2 RT-PCR. The horizontal black line is the antigen test positivity cutoff, corresponding to the average plus four standard deviations of the signals induced by SARS-CoV-2 RT-PCR negative samples. The vertical black line separates SARS-CoV-2 RT-PCR positive (n = 48) and negative (n = 96) NPS samples. The coloring in the graphs indicates presence (red) or absence (blue) of cytopathic effect (CPE) following inoculation of VE6-TMPRSS2-H10 cells with 50 µl of the NPS sample, black = not cultured. TR-FRET = time-resolved Förster resonance energy transfer, NP = nucleoprotein, RBD = receptor-binding domain, HTRF = homogeneous time-resolved fluorescence.

### False positivity and cross-reactivity

After finding a total antibody concentration of 12 nM ideal for the assay performance, we were interested in knowing the rate of false positive results. To that end, we tested in the TR-FRET antigen assays 96 SARS-CoV-2 RT-PCR negative NPS samples. In parallel, we studied the potential cross-reactivity of cell culture grown seasonal common cold coronaviruses hCoV-229E and hCoV-NL63 in the TR-FRET assays. Among the SARS-CoV-2 RT-PCR negative NPS samples, only one produced a positive HTRF signal. We reanalyzed this sample with another RT-PCR assay (Xpert Xpress SARS-CoV-2, Cepheid), confirming the negative result. To assess the antigen-specificity of the signal, we also analyzed this sample using mismatching combinations of the labeled anti-NP and anti-RBD antibodies. All of the combinations yielded a positive result, suggesting that something other than the antigens bring the labeled antibodies together. By using the optimized conditions with 12 nM total labeled antibodies, the specificity of NP and SP TR-FRET assays in comparison with RT-PCR were 99.0% (95/96) and in comparison with virus isolation 100% (10/10). We then set as cutoffs for the TR-FRET assays the average plus four standard deviations of the signals from the SARS-CoV-2 RT-PCR negative NPS samples (excluding the single outlier). Using these cutoffs neither hCoV-229E nor hCoV-NL63 yielded a significant TR-FRET signal in either NP or SP assay (Fig. 3). The results with the cutoff values selected using the negative NPS samples concurred with those by the arbitrary cutoffs utilized earlier, and are summarized in Fig. 2.

**Figure 3.**
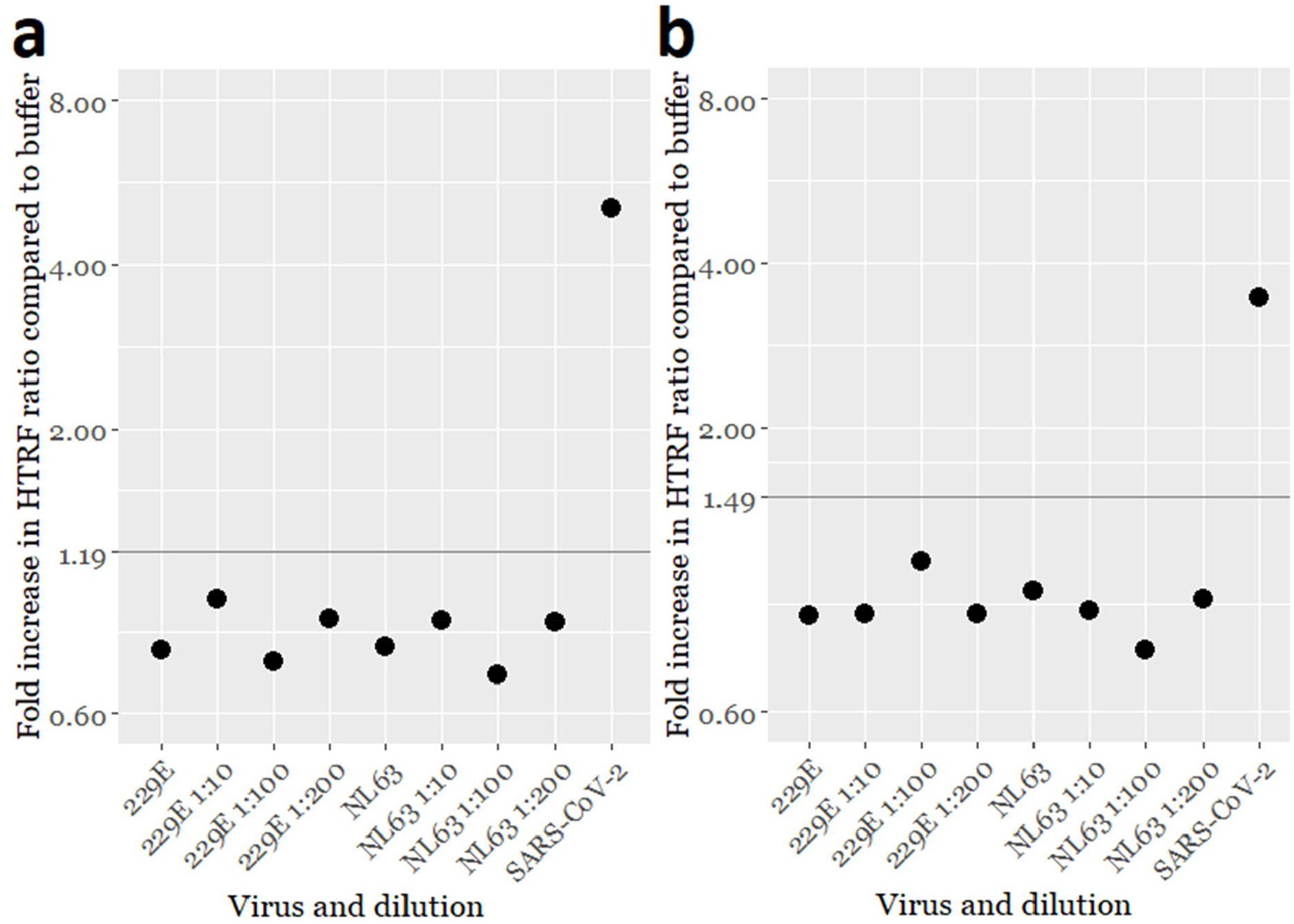
SARS-CoV-2 TR-FRET antigen assay cross-reactivity evaluated with cell culture supernatants of seasonal human coronaviruses hCoV-229E and -NL63. a) Anti-NP assay results with HCoV-229E and -NL63 cell culture supernatants at different dilutions. b) Anti-NP assay results with HCoV-229E and -NL63 cell culture supernatants at different dilutions. The y-axis (log scale) indicates a fold increase in HTRF ratio (HTRF_sample_/HTRF_buffer_). The horizontal lines indicate the respective TR-FRET assay cutoffs. UV-inactivated SARS-CoV-2 containing cell culture supernatant is included as a positive control.

## DISCUSSION

The SARS-CoV-2 epidemic that began in China late 2019 quickly evolved into pandemic in the spring of 2020. After an initial lag in ramping up the testing capacity, RT-PCR quickly became the gold standard of acute SARS-CoV-2 diagnostics. While RT-PCR is very sensitive in picking up individuals with acute infection, the downside is that patients recovering from COVID-19 can remain RT-PCR positive over a long period. Antigen testing on the other hand is somewhat less sensitive for detecting the patients with acute infection; however, there appears to be a better correlation between presence of antigen and infectious virus in the NPS samples. In this manuscript, we describe a laboratory-developed test for the detection of SARS-CoV-2 antigen in NPS samples and compare the test’s performance against virus isolation and RT-PCR. The assay is quick and very easy to use, furthermore the assay is rather uncomplicated to set up provided that specific antibodies against the structural proteins of the virus are available. While we employ polyclonal antibodies in the assay, it likely could be set up using monoclonals. The fluorophores (chelated Eu and AlexaFluor 647) are readily available, and the results can be read on any microplate reader capable of measuring time-resolved fluorescence. Notably, we set up the assay in detergent-containing RIPA buffer, which contains 1% NP-40 and 0.1% SDS, both of which inactivate SARS-CoV-2^21,22^. Thus collecting the NPS samples directly into this matrix would significantly increasing the end user safety.

We set up the assay for the detection of both NP and SP of SARS-CoV-2 but observed a clear difference between the LODs of the two assays with NP being detected at approximately 35 times lower concentration. This is likely explained by the fact that we employed an antibody directed against the RBD, which constitutes only about one sixth of SP. The fact that the antibody in use recognizes only a single domain could make it sterically impossible for two antibody molecules to bind a single SP molecule, and thus we speculate the obtained signal to come from SP trimers i.e. spikes. NP is more abundant in both virions and infected cells (refer to Fig. S7), which additionally contributed to the higher sensitivity of NP detection in cell culture supernatants and NPS samples.

Performance analysis of the antigen assay using NPS samples from 48 SARS-CoV-2 RT-PCR positive and 96 negative individuals revealed that the NP assay correctly identified 37 of the positive samples, and all but one of the negatives. All the 37 true positives had a Ct value <25 cycles in the diagnostic RT-PCR. Similar to other studies, we observed a strong association between the sample infectivity and positive antigen test result; of the 38 samples that yielded an isolate 37 produced a positive result in the NP assay. We used 50 µl for the virus isolation while the antigen assay only takes 10 µl, which could explain why one of the samples with infectious virus was not picked up. We intentionally selected NPS samples over a broad range of Ct values in SARS-CoV-2 RT-PCR to obtain an estimate of the detection limit as compared to RT-PCR. The fact that we analyzed samples that were not collected fresh and had been subjected to at least one freeze-thaw cycle may have negatively affected the assay sensitivity. In any case, our test could detect 97.4% of the NPS samples with infectious virus.

Because both NP and SP antigen assays gave a positive result for a single SARS-CoV-2 RT-PCR negative sample, we re-analyzed it using a different RT-PCR with a negative result. We also attempted virus isolation from the sample but without success. It is possible that the false positive result is a result of cross-reactivity to a human coronavirus, hCoV, infection. Of the hCoVs only hCoV-NL63 and SARS-CoV use the same receptor as SARS-CoV-2, i.e. the angiotensin converting enzyme 2 (ACE2)^24,25^. Thus it would be most logical that the reactivity of this sample would be due to hCoV-NL63 because both SP (based on RBD) and NP assays gave a positive result. We did however test the two assays using cell culture -grown hCoV-229E and hCoV-NL63, yet with negative results. It appears that the sample yielding a false positive reaction contained an interfering substance, which caused the immunoglobulins to aggregate, because also labeled antibodies against different antigens yielded a TR-FRET signal. The use of miss-matching antibodies could be in the future serve to discern true and false positives in the TR-FRET assay.

In conclusion, we report the development of a rapid SARS-CoV-2 antigen test based on simultaneous binding of two or more fluorophore-labeled antibody molecules to the antigen. The TR-FRET –based antigen test is rapid to perform, and its results correlate well with the presence of infectious virus in clinical sample. The assay is easy to set up, if a suitable antibody against SARS-CoV-2 NP and a plate reader enabling TR-FRET measurement are available. We estimate that a single plate reader and an experienced technician could manually analyze hundreds of specimens per hour, ideally with a 30-minute turn-around time from sample arrival to results (see supplementary information for details). The assay throughput could significantly be up scaled by using automation, and alike RT-PCR the sample collection would represent the major limiting factor. The NPS sampling directly to a detergent-containing buffer increases the assay’s user-safety. We envision that the assay could be applied widely in the field, e.g. hospitals, retirement homes, airports and train stations, and in schools to identify people likely to spread the virus.

## Data Availability

The data referred to in the manuscript is either provided as supplementary material or upon request.

## ACKNOWLEDGEMENTS

The authors wish to acknowledge Drs. Sisko Tauriainen (University of Turku, Turku, Finland) and Lia van der Hoek (Academic Medical Center, Amsterdam, Netherlands) for providing the human coronaviruses used for evaluating cross-reactivity. The study was supported by Academy of Finland (grants 308613, 314119, and 335762 to JH).

## SUPPLEMENTARY INFORMATION

### Assay throughput estimate

Using 384-well plates, 192 samples can be measured in duplicate on one plate. Pipetting the reagent mixes and samples manually on plate with a multichannel pipette is estimated to require 10 minutes for a full plate (2-2.5 minutes for reagent mixes and 7.5-8 minutes for samples, longer for the latter due to tip change between samples). Measuring the HTRF ratio in triplicate takes approximately 18 min for a full 384-well plate using Hidex Sense (Hidex Oy, Finland). HTRF ratios are calculated automatically and the results can be exported as e.g. an Excel file. While the measurement takes place, the next plate can be prepared. With measurement intervals of 20 minutes, a single machine and one operator could ideally measure 576 samples/hour that would correspond to 13 824 samples/24 hours, assuming shift work. The throughput could be increased by including automation and additional staff to aid with sample preparation and the transfer of results to a laboratory information management system.

## Supplementary figures and tables

**Figure S1.**
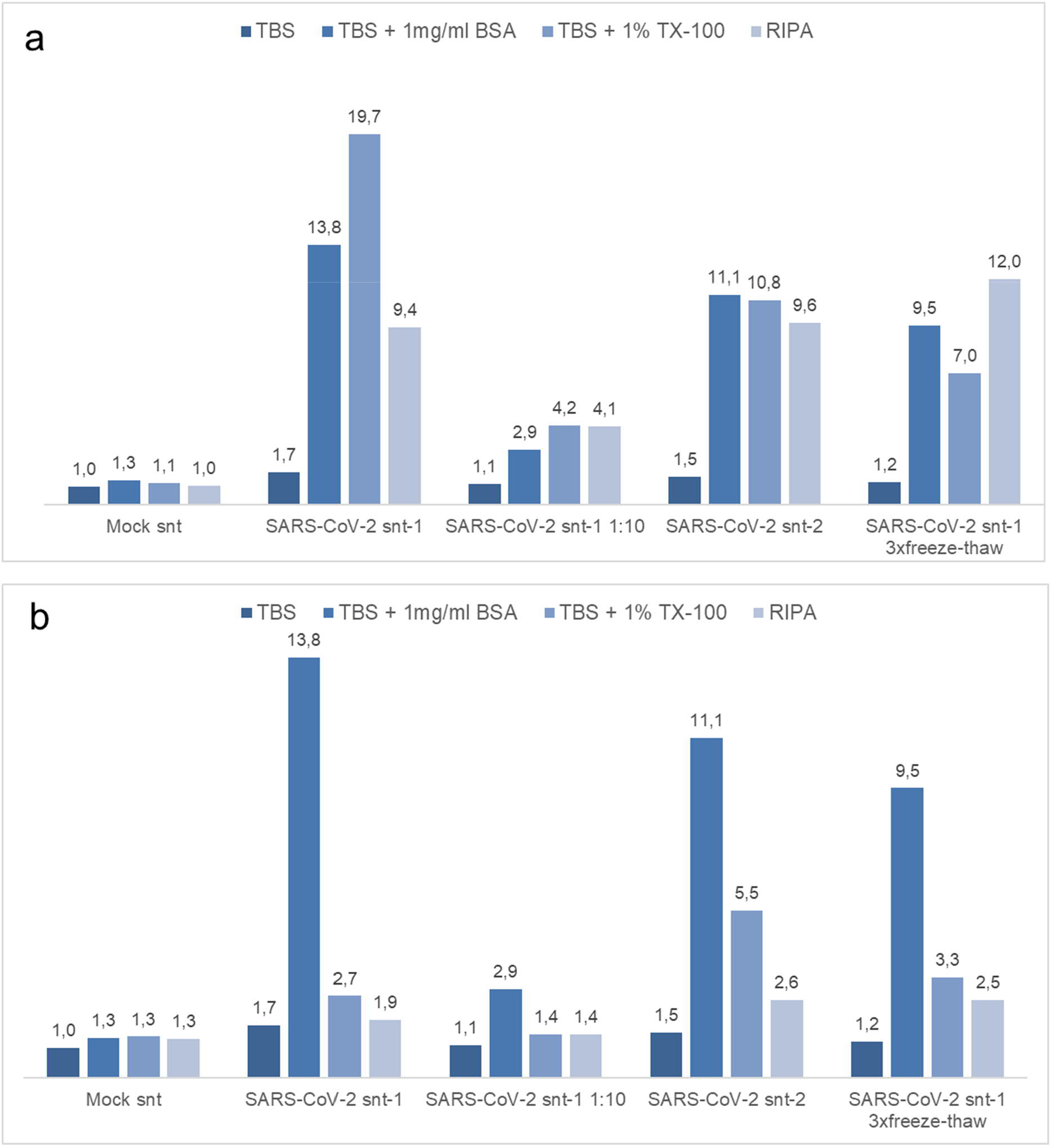
TR-FRET antigen assay in different buffer compositions. The antibody concentrations refer to the total amount used in reaction i.e. a 1:1 mixture of Eu- and AF647-labeled antibodies. a) Anti-NP at 1 µM concentration. b) Anti-NP at 1 µM concentration. The y-axis (and values over bars) indicates a fold increase in HTRF ratio (HTRF_sample_/HTRF_buffer_). Mock snt = supernatant from mock-infected Vero E6 cells, SARS-CoV-2 snt-1 and −2 = supernatants containing infectious SARS-CoV-2.

**Figure S2.**
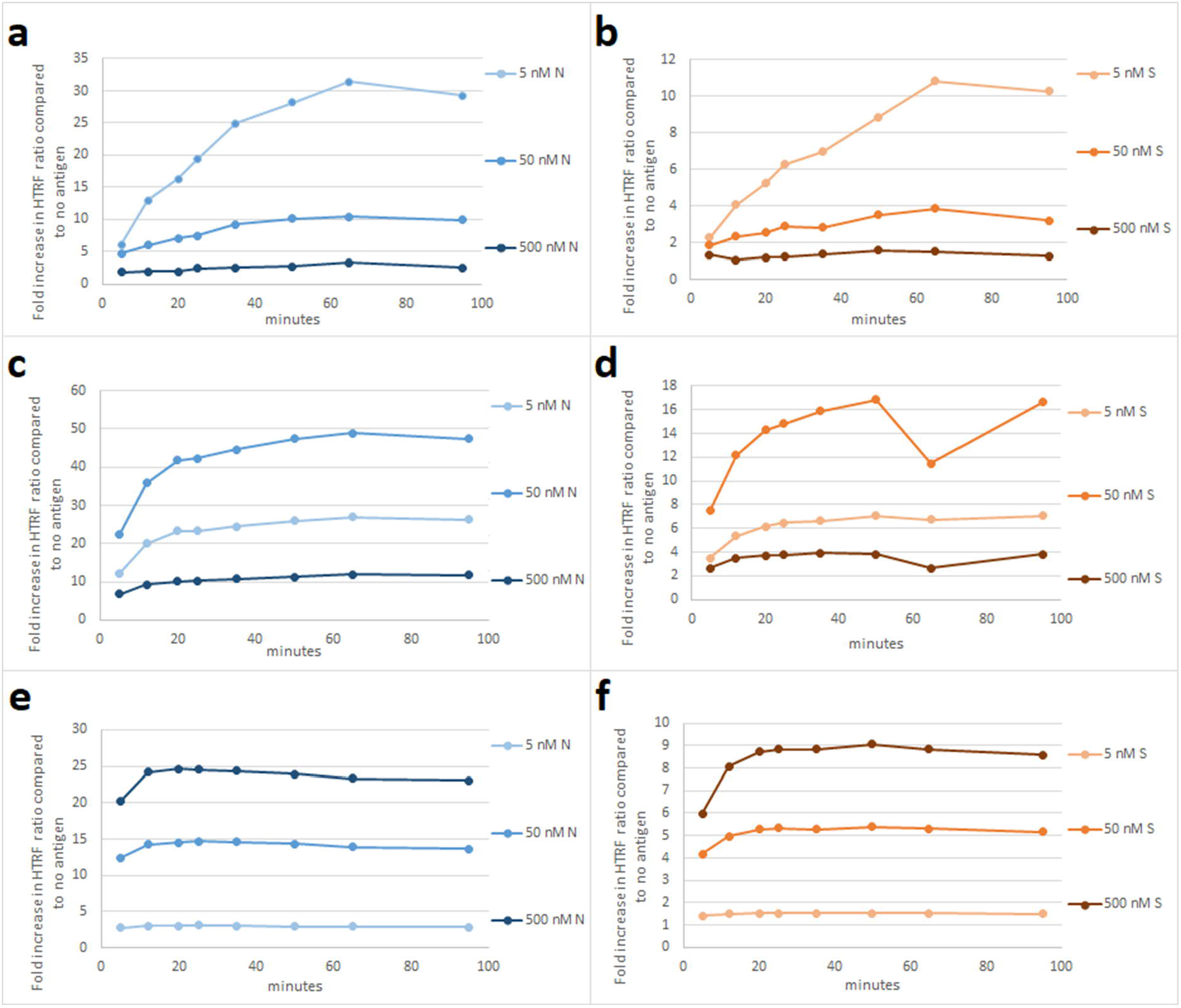
TR-FRET cross-titration of labeled antibodies and purified recombinant NP and SP. a) Anti-NP antibody concentration of 5 nM against NP concentrations of 5, 50 and 500 nM. b) Anti-RBD antibody concentration of 5 nM against SP concentrations of 5, 50 and 500 nM. c) Anti-NP antibody concentration of 50 nM against NP concentrations of 5, 50 and 500 nM. d) Anti-RBD antibody concentration of 50 nM against SP concentrations of 5, 50 and 500 nM. e) Anti-NP antibody concentration of 500 nM against NP concentrations of 5, 50 and 500 nM. f) Anti-RBD antibody concentration of 500 nM against SP concentrations of 5, 50 and 500 nM. The antibody concentrations refer to the total amount used in reaction i.e. a 1:1 mixture of Eu- and AF647-labeled antibodies. The y-axis indicates a fold increase in HTRF ratio (HTRF_sample_/HTRF_buffer_). The x-axis shows time in minutes since the pipetting of the samples on plate began (∼5 minutes before the first measurement). NP = nucleoprotein, SP = spike glycoprotein, RBD = receptor-binding domain.

**Table S1.** Comparison of TR-FRET antigen assay results, expressed as HTRF ratio (HTRF_sample_/HTRF_buffer_), using different ratios of Eu- and AF647-labeled anti-NP and anti-RBD antibodies. NP = nucleoprotein, RBD = receptor-binding domain, Eu = Eu-labeled, AF = Alexa Fluor 647 labeled.

| HTRF ratio compared to average of mock media 1:1 and 1:100 |  |  |  |  |  |  |  |  |
| --- | --- | --- | --- | --- | --- | --- | --- | --- |
|  | 10 µl VE6 |  |  |  |  |  | 10 µl VE6 mock-media |  |
|  | 1:1 | 1:10 | 1:50 | 1:100 | 1:1000 |  | 1:1 | 1:100 |
| anti-NP 10 nM Eu + 10 nM AF | 18.8 | 15.9 | 4.4 | 2.7 | 1.2 |  | 1.0 | 1.0 |
| anti-NP 20 nM Eu + 20 nM AF | 26.1 | 12.8 | 3.7 | 2.5 | 1.2 |  | 1.0 | 1.0 |
| anti-NP 40 nM Eu + 40 nM AF | 45.0 | 10.4 | 2.9 | 2.1 | 1.1 |  | 1.0 | 1.0 |
| anti-NP 80 nM Eu + 80 nM AF | 39.5 | 7.0 | 2.2 | 1.7 | 1.1 |  | 1.0 | 1.0 |
| anti-NP 13 nM Eu + 27 nM AF | 23.0 | 16.5 | 4.5 | 2.8 | 1.2 |  | 1.0 | 1.0 |
| anti-NP 27 nM Eu + 13 nM AF | 13.0 | 14.9 | 4.5 | 2.8 | 1.2 |  | 1.0 | 1.0 |
| anti-RBD 10 nM Eu + 10 nM AF | 3.6 | 1.3 | 1.3 | 1.1 | 1.1 |  | 1.0 | 1.0 |
| anti-RBD 20 nM Eu + 20 nM AF | 3.3 | 1.3 | 1.1 | 1.0 | 1.0 |  | 1.0 | 1.0 |
| anti-RBD 40 nM Eu + 40 nM AF | 2.7 | 1.2 | 1.1 | 1.0 | 1.0 |  | 1.0 | 1.0 |
| anti-RBD 80 nM Eu + 80 nM AF | 2.3 | 1.2 | 1.0 | 1.0 | 1.0 |  | 1.0 | 1.0 |
| anti-RBD 13 nM Eu + 27 nM AF | 3.1 | 1.2 | 1.0 | 1.0 | 0.9 |  | 1.0 | 1.0 |
| anti-RBD 27 nM Eu + 13 nM AF | 2.3 | 1.1 | 1.0 | 1.1 | 1.0 |  | 1.0 | 1.0 |

**Figure S3.**
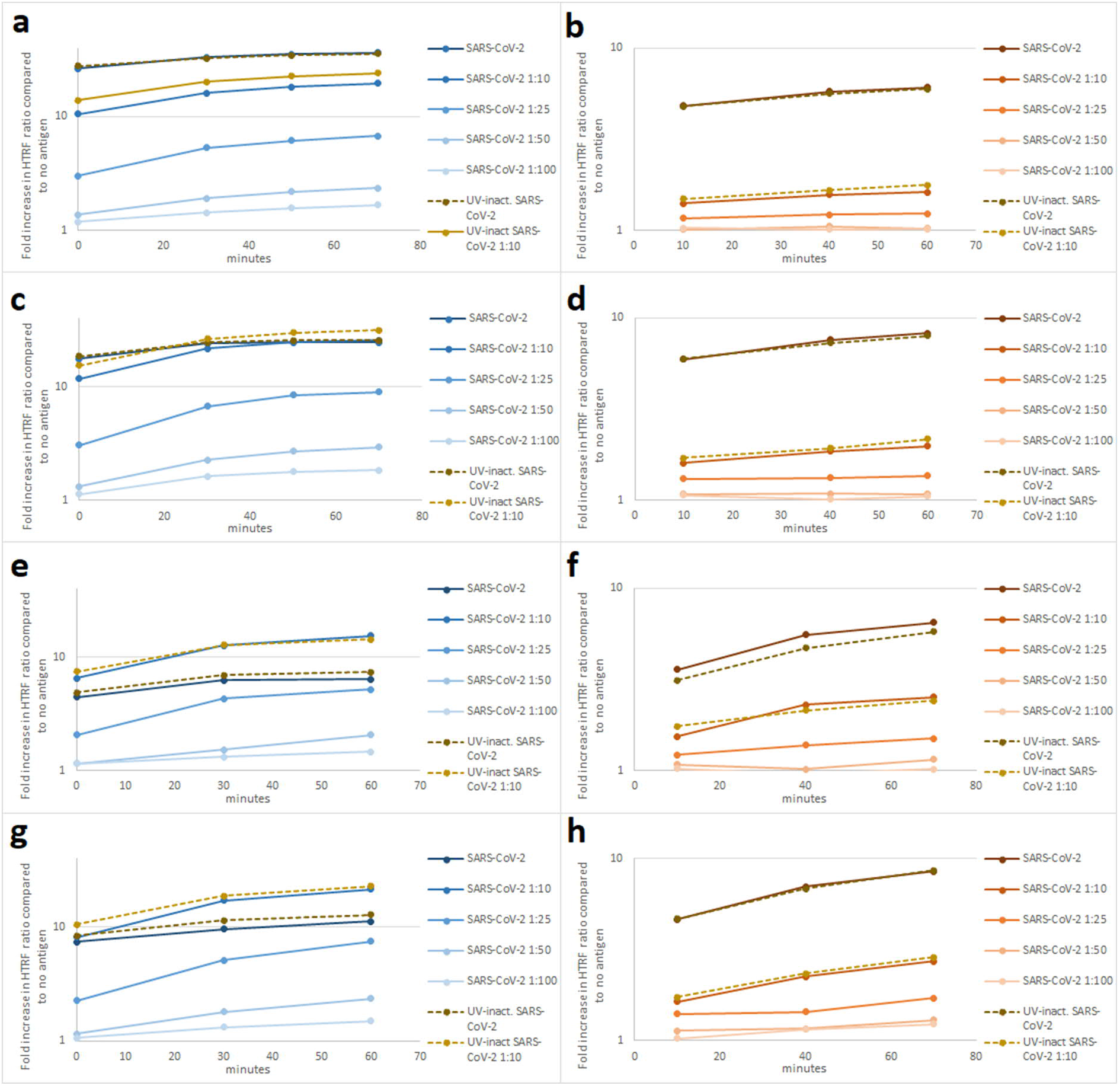
TR-FRET cross-titration of labeled antibodies and virus-containing cell culture supernatants. The antibody concentrations refer to the total amount used in reaction i.e. a 1:1 mixture of Eu- and AF647-labeled antibodies. The different antibody concentrations were titrated against SARS-CoV-2 containing cell culture supernatant spiked at varying dilutions in a pool of negative NPS samples. a) Anti-NP antibody at 50 nM. b) Anti-RBD antibody at 50 nM. c) Anti-NP antibody at 25 nM. d) Anti-RBD antibody at 25 nM. e) Anti-NP antibody at 12 nM. f) Anti-RBD antibody at 12 nM. g) Anti-NP antibody at 6 nM. d) Anti-RBD antibody at 6 nM. The y-axis (log scale) indicates a fold increase in HTRF ratio (HTRF_sample_/HTRF_buffer_). The x-axis shows time in minutes since the first measurement began.

**Figure S4.**
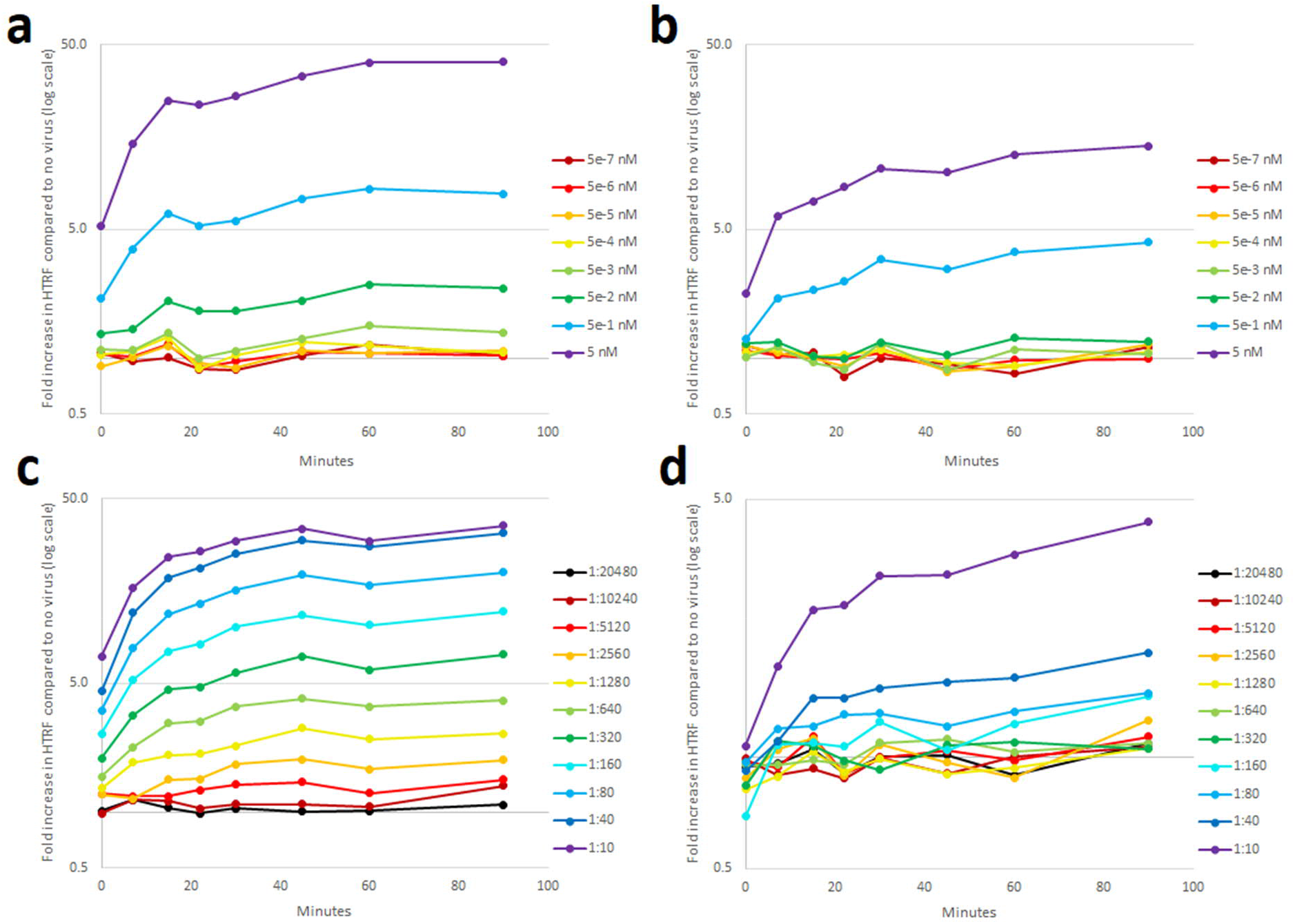
The limit of detection for TR-FRET antigen detection using NPS samples spiked with recombinant antigens or inactivated virus. The evaluation is with total antibody concentrations of 12 nM, i.e. 6 nM of Eu- and 6 nM of AF647-labeled antibodies. a) Recombinant NP spiked at 0.5 fM to 5 nM in a pool of negative NPS samples. b) Recombinant SP spiked at 0.5 fM to 5 nM in a pool of negative NPS samples. c) UV inactivated SARS-CoV-2 containing cell culture supernatant spiked at 1:20480 to 1:10 in a negative NP swab sample, with labeled anti-N antibodies at 6+6 nM. d) Inactivated SARS-CoV-2 spiked at 1:20480 to 1:10 in a pool of negative NPS samples. The y-axis (log scale) indicates a fold increase in HTRF ratio (HTRF_sample_/HTRF_buffer_). The x-axis shows time in minutes since the first measurement began. NP = nucleoprotein, SP = spike glycoprotein, RBD = receptor-binding domain.

**Figure S5.**
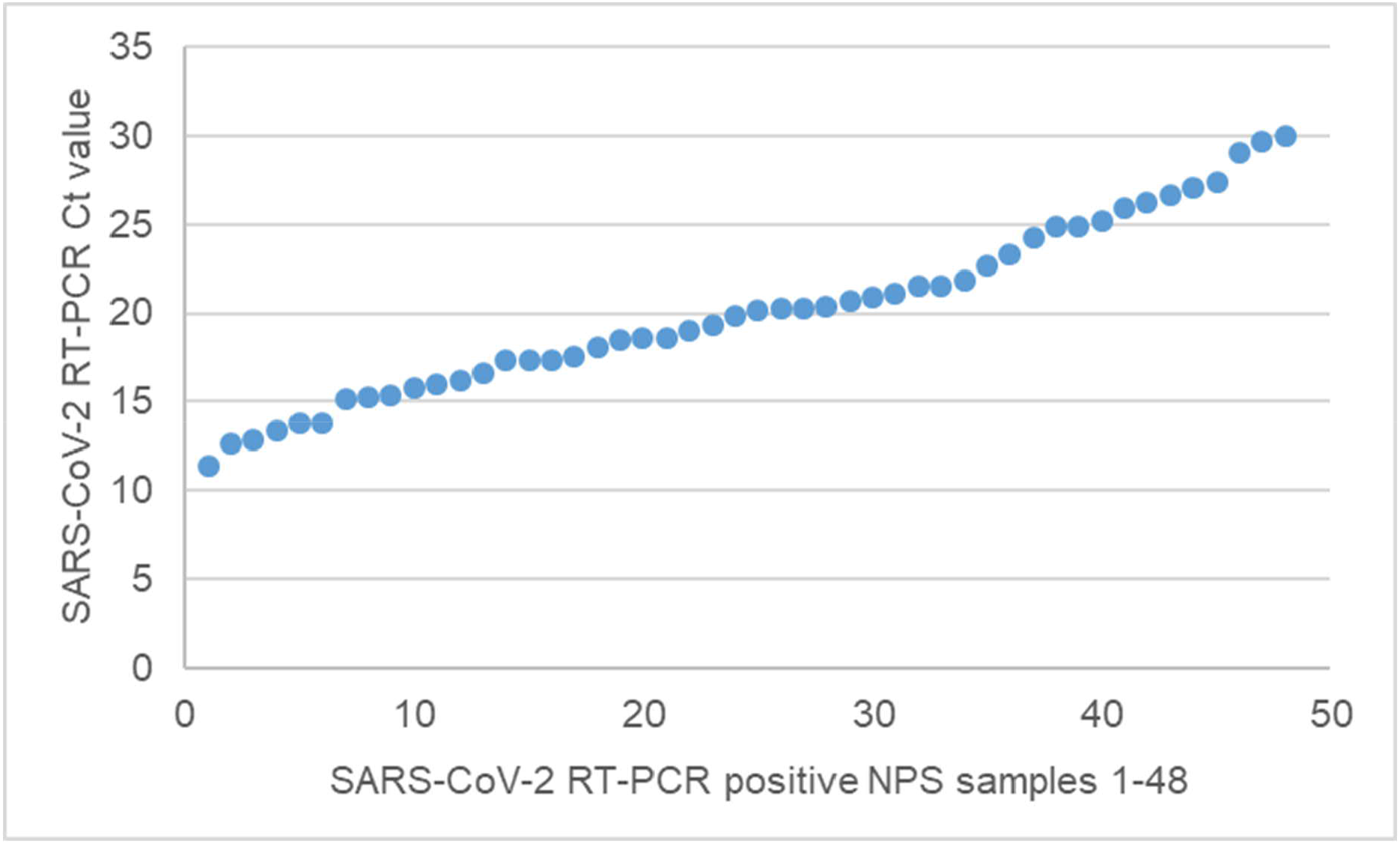
Distribution of the Ct values in the original diagnostic SARS-CoV-2 RT-PCR. The y-axis shows the Ct value in the RT-PCR and X-axis shows the sample number.

**Figure S6.**
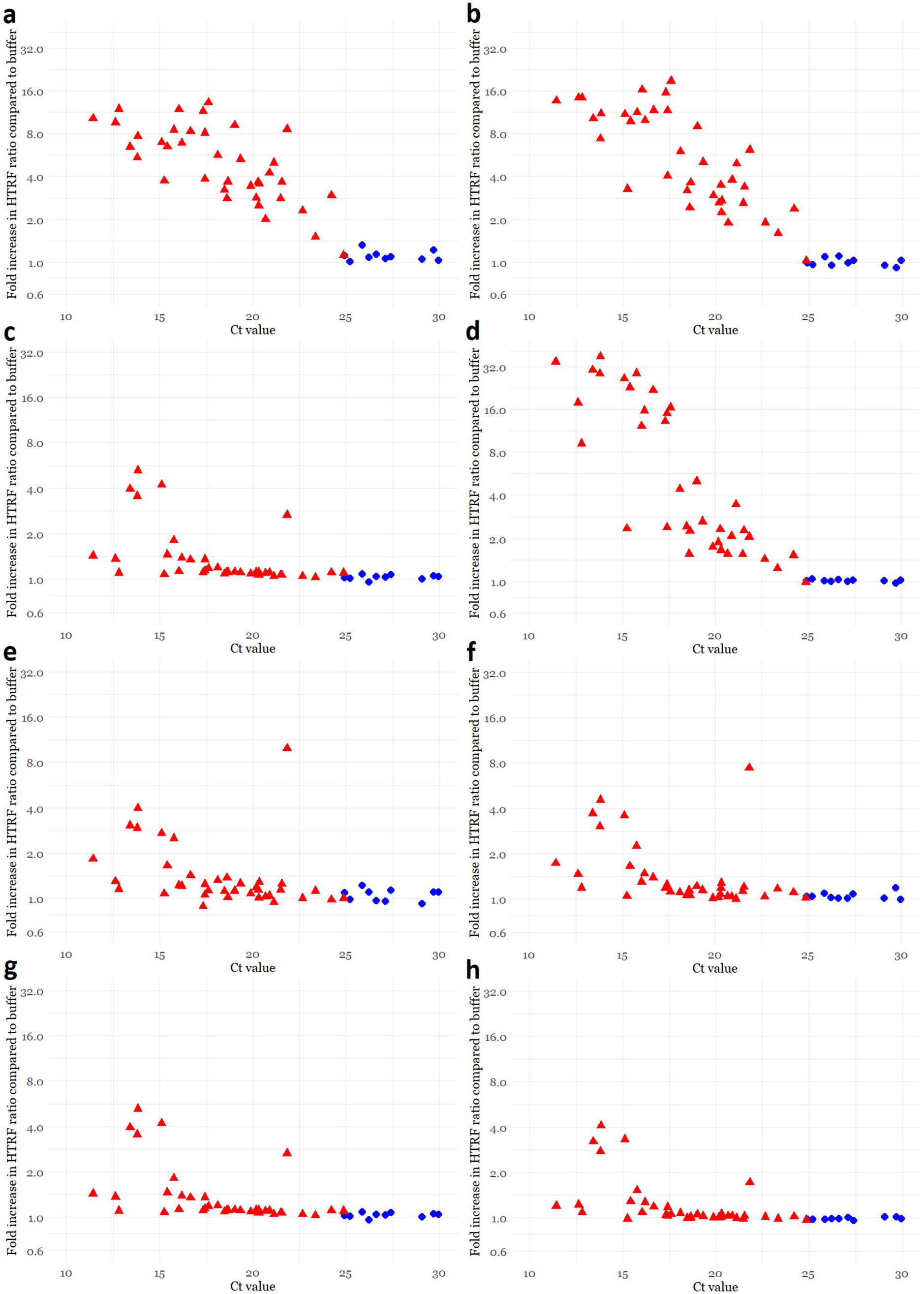
TR-FRET antigen assay results for NPS samples at varying antibody concentrations (6 to 50 nM) against Ct value of the positive SARS-CoV-2 RT-PCR result. a) Anti-NP antibody at 6 nM concentration. b) Anti-NP antibody at 12 nM concentration. c) 1Anti-NP antibody at 25 nM concentration. d) Anti-NP antibody at 50 nM concentration. e) Anti-RBD antibody at 6 nM concentration. f) Anti-RBD antibody at 12 nM concentration. g) Anti-RBD antibody at 25 nM concentration. h) Anti-RBD antibody at 50 nM concentration. The y-axis (log scale) indicates a fold increase in HTRF ratio (HTRF_sample_/HTRF_buffer_) measured directly after pipetting of the samples on plate. The x-axis shows the Ct value measured in the diagnostic SARS-CoV-2 RT-PCR. The coloring in the graphs indicates presence (red) or absence (blue) of cytopathic effect (CPE) following inoculation of VE6-TMPRSS2-H10 cells with 50 µl of the NPS sample. FRET = Förster resonance energy transfer, NP = nucleoprotein, RBD = receptor-binding domain, Ct = cycle threshold, Eu = europium, AF = Alexa Fluor 647.

**Figure S7.**
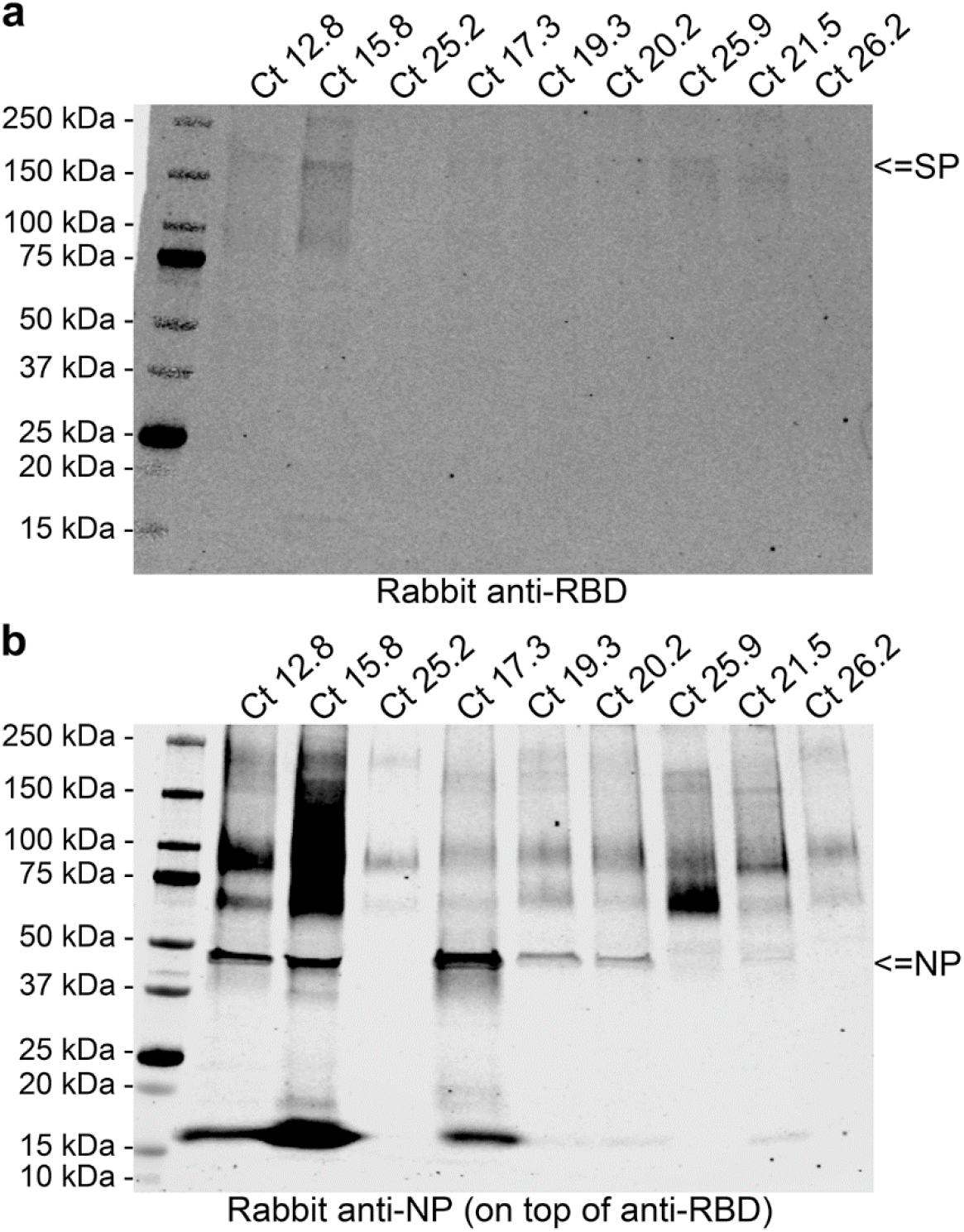
Detection of SARS-CoV-2 antigens in selected NPS samples by immunoblotting. The NPS samples selected based on the Ct values in the diagnostic SARS-CoV-2 RT-PCR were separated on SDS-PAGE (4-15% Mini-PROTEAN TGX Precast Protein Gel, deep well, Bio-RAD), approximately 40 µl of NPS sample/well, using standard protocol but under non-reducing conditions. The proteins transferred (standard wet blotting protocol) onto nitrocellulose membrane (Nitrocellulose Blotting Membrane, Amersham Protran 0.45 µm NC, GE Healthcare) were following blocking (30 min at room temperature, blocking buffer: 3% skimmed milk powder in 50 mM Tris, 150 mM NaCl, 0.05% Tween 20) first probed with anti-RBD antiserum (1:3000 diluted in blocking buffer), and the binding detected using IRDye 800CW Donkey anti-rabbit IgG secondary antibody (LI-COR Biosciences), 1:10000 dilution in blocking buffer. The membrane was then probed with anti-NP antiserum (1:4000 diluted in blocking buffer), and the same secondary antibody was used for the detection. The results were recorded using Odyssey Infrared Imaging System (LI-COR Biosciences). **a)** The result of anti-RBD (detects S protein under non-reducing conditions) staining. The wells are labeled by the diagnostic SARS-CoV-2 RT-PCR values of the NPS samples. A band migrating at approximately 170 kDa (indicated by an arrow) represents the unprocessed SP and it is detected in some of the samples. **b)** The result of anti-NP staining. The labeling of wells as in **a**. A band migrating at approximately 45 kDa (indicated by an arrow) represents the NP and it is clearly detectable in samples with Ct values <21.5.

**Figure S8.**
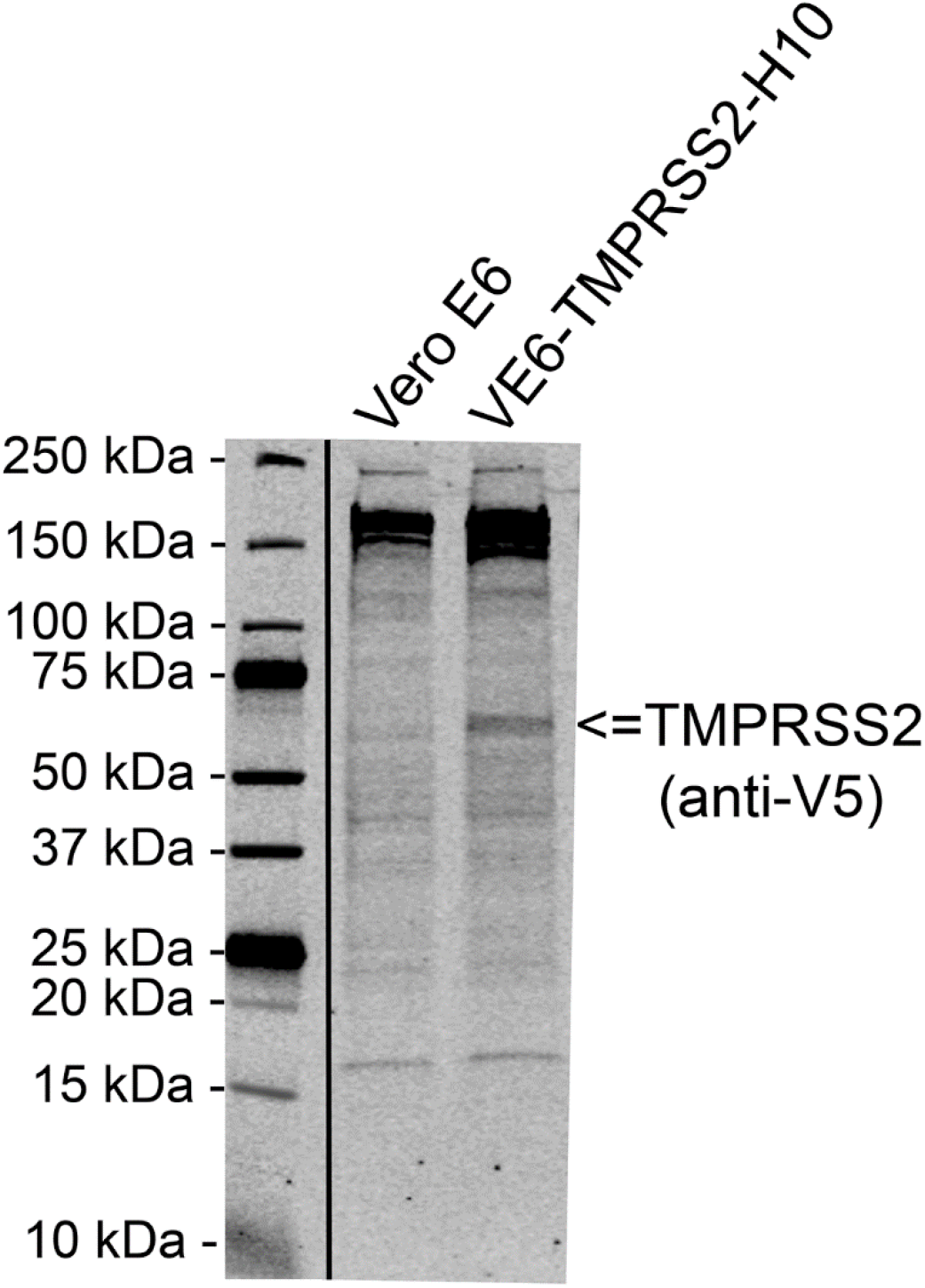
Detection of Transmembrane serine protease 2 (TMPRSS2) in the clonal population of lentivirus-transduced Vero E6 cells expressing TMPRSS2 (VE6-TMPRSS2-H10). Wild-type Vero E6 and VE6-TMPRSS2-H10 cells were collected from a 6-well plate using cell scraper, washed twice with PBS, and lyzed in Laemmli Sample Buffer. The cell lysates were were separated on SDS-PAGE (4-20% Mini-PROTEAN TGX Precast Protein Gel, Bio-RAD), using standard protocol. The proteins transferred (standard wet blotting protocol) onto nitrellulose membrane (Nitrocellulose Blotting Membrane, Amersham Protran 0.45 µm NC, GE Healthcare) were following blocking (30 min at room temperature, blocking buffer: 3% skimmed milk powder in 50 mM Tris, 150 mM NaCl, 0.05% Tween 20) probed with mouse monoclonal anti-V5 epitope tag antibody (clone E10/V4RR, Invitrogen), diluted 1:2000 in blocking buffer. The binding was detected using IRDye 800CW Donkey anti-mouse IgG secondary antibody (LI-COR Biosciences), 1:10000 dilution in blocking buffer, and the results were recorded using Odyssey Infrared Imaging System (LI-COR Biosciences). The band indicated by an arrow at approximately 60-65 kDa by migration indicates the expression of V5-tagged TMPRSS2 by the VE6-TMPRSS2-H10 clone of Vero E6.

**Figure S9.**
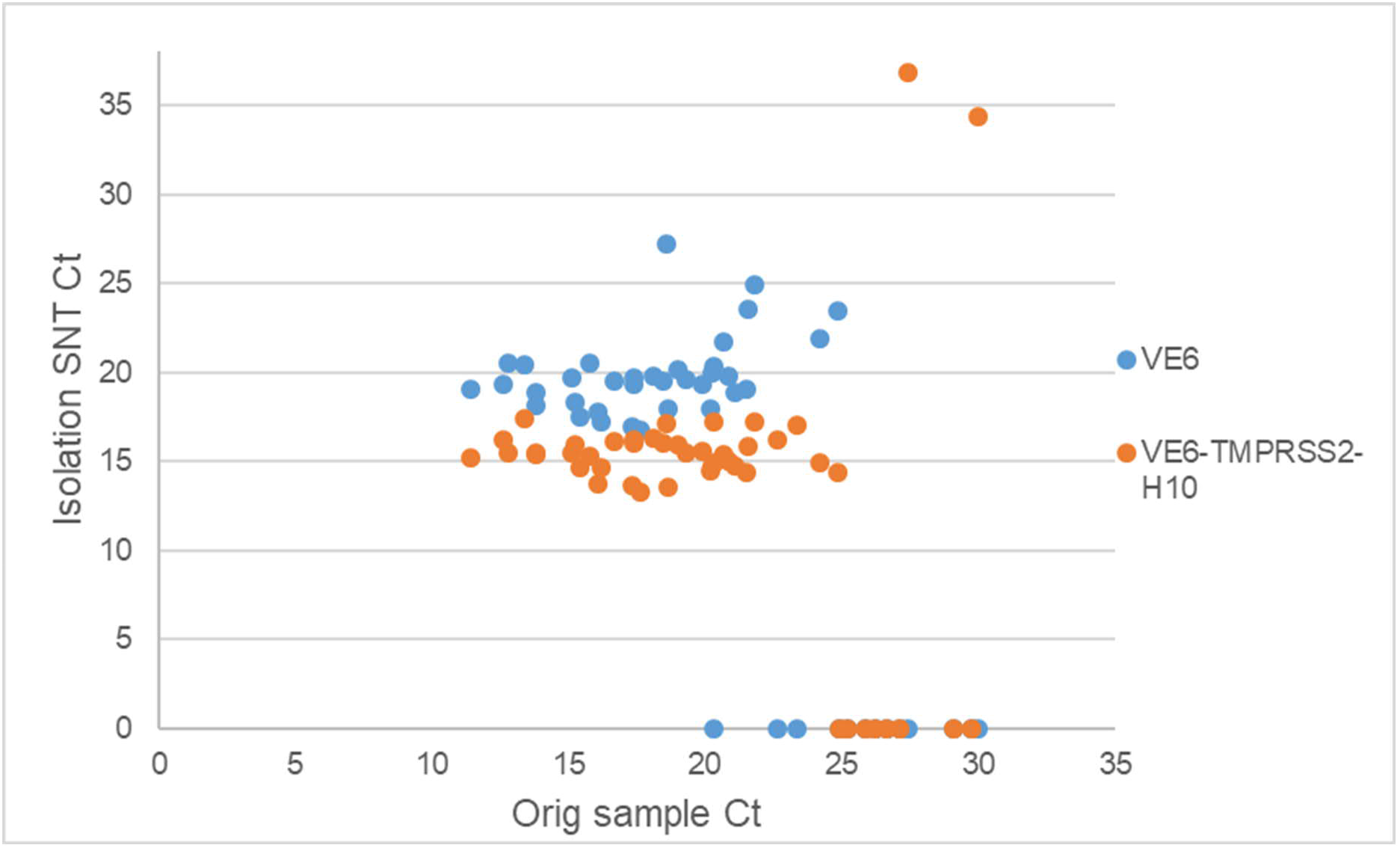
RT-PCR results from supernatants collected at 5 days post inoculation with SARS-CoV-2 RT-PCR positive NPS samples. Y-axis shows Ct value from SARS-CoV-2 RT-PCR from the cell culture supernatant and X-axis shows the respective Ct value of the original diagnostic test. The blue dots indicate supernatant from isolation attempt on Vero E6 cells and the orange dots indicate supernatant from isolation attempt on VE6-TMRPRSS2-H10 cells.

## REFERENCES

1) Arons M.M. et al. Presymptomatic SARS-CoV-2 infections and transmission in a skilled nursing facility. N Engl J Med 382, 2081–2090 (2020). https://doi:10.1056/NEJMoa2008457.

2) Bullard J. et al. Predicting Infectious SARS-CoV-2 From Diagnostic Samples. Clin Infect Dis. (2020) https://doi.org/10.1093/cid/ciaa638.

3) Wölfel R., et al. Virological assessment of hospitalized patients with COVID-2019. Nature 58, 465–469 (2020) https://doi.org/10.1038/s41586-020-2196-x.

4) Pekosz A., et al. Antigen-based testing but not real-time PCR correlates with SARS-CoV- 2 virus culture. medRxiv (2020). https://doi.org/10.1101/2020.10.02.20205708.

5) Mina M.J., et al. Rethinking Covid-19 Test Sensitivity - A Strategy for Containment. N Engl J Med 383, e120 (2020). https://doi.org/10.1056/NEJMp2025631.

6) Dinnes J., et al. Rapid, point-of-care antigen and molecular-based tests for diagnosis of SARS-CoV-2 infection. Cochrane Database Syst Rev CD013705 (2020). https://doi.org/10.1002/14651858.CD013705.

7) U.S. Food and Drug Administration. Coronavirus Disease 2019 (COVID-19) Emergency Use Authorizations for Medical Devices: In Vitro Diagnostic EUAs. Updated 4.12.2020. Accessed 5.12.2020. https://www.fda.gov/medical-devices/coronavirus-disease-2019-covid-19-emergency-use-authorizations-medical-devices/vitro-diagnostics-euas.

8) Saraheimo S., et al. Time-resolved FRET -based approach for antibody detection - a new serodiagnostic concept. PLOS One 8, e62739 (2013) https://doi.org/10.1371/journal.pone.0062739.

9) Hepojoki S., et al. A protein L-based immunodiagnostic approach utilizing time-resolved Förster resonance energy transfer. PLOS One 9, e106432 (2014). https://doi.org/10.1371/journal.pone.0106432.

10) Hepojoki S., et al. Rapid homogeneous immunoassay based on time-resolved Förster resonance energy transfer for serodiagnosis of acute hantavirus infection. J Clin Microbiol 53, 636–640 (2015). https://doi.org/10.1128/JCM.02994-14.

11) Hepojoki S., et al. Competitive Homogeneous Immunoassay for Rapid Serodiagnosis of Hantavirus Disease. J Clin Microbiol 53, 2292–2297 (2015). https://doi.org/10.1128/JCM.00663-15.

12) Kareinen L., et al. Immunoassay for serodiagnosis of Zika virus infection based on time- resolved Förster resonance energy transfer. PLOS One 14, e0219474 (2019) https://doi.org/10.1371/journal.pone.0219474.

13) Rusanen J., et al. LFRET, a novel rapid assay for anti-tissue transglutaminase antibody detection. PLOS One 14, e0225851 (2019). https://doi.org/10.1371/journal.pone.0225851.

14) Rusanen J., et al. Rapid homogeneous assay for detecting antibodies against SARS-CoV- 2. medRxiv (2020). https://doi.org/10.1101/2020.11.01.20224113.

15) Mannonen L., et al. Comparison of two commercial platforms and a laboratory developed test for detection of SARS-CoV-2 RNA. medRxiv (2020). https://doi.org/10.1101/2020.07.03.20144758.

16) Corman V.M., et al. Detection of 2019 novel coronavirus (2019-nCoV) by real-time RT- PCR. Euro Surveill 25, 2000045 (2020). https://doi.org/10.2807/1560-7917.ES.2020.25.3.2000045.

17) Berthoux L., et al. Lv1 Inhibition of Human Immunodeficiency Virus Type 1 Is Counteracted by Factors That Stimulate Synthesis or Nuclear Translocation of Viral cDNA. J Virol 78, 11739–50 (2004). https://doi.org/10.1128/JVI.78.21.11739-11750.2004.

18) Haveri A., et al. Serological and molecular findings during SARS-CoV-2 infection: the first case study in Finland, January to February 2020. Euro Surveill 25, 2000266 (2020). https://doi.org/10.2807/1560-7917.ES.2020.25.11.2000266.

19) Stadlbauer D., et al. SARS-CoV-2 Seroconversion in Humans: A Detailed Protocol for a Serological Assay, Antigen Production, and Test Setup. Curr Protoc Microbiol 57, e100 (2020). https://doi.org/10.1002/cpmc.100.

20) Amanat F., et al. A serological assay to detect SARS-CoV-2 seroconversion in humans. Nat Med 26, 1033–1036 (2020). https://doi.org/10.1038/s41591-020-0913-5.

21) Welch S.R., et al. Analysis of Inactivation of SARS-CoV-2 by Specimen Transport Media, Nucleic Acid Extraction Reagents, Detergents, and Fixatives. J Clin Microbiol (2020) https://doi.org/10.1128/JCM.01713-20.

22) Patterson E.I., et al. Methods of Inactivation of SARS-CoV-2 for Downstream Biological Assays. J Infect Dis (2020). https://doi.org/10.1093/infdis/jiaa507.

23) Hoffman M., et al. SARS-CoV-2 Cell Entry Depends on ACE2 and TMPRSS2 and Is Blocked by a Clinically Proven Protease Inhibitor. Cell 181, 271-280.e8 (2020). https://doi.org/10.1016/j.cell.2020.02.052.

24) Hofmann H., et al. Human coronavirus NL63 employs the severe acute respiratory syndrome coronavirus receptor for cellular entry. Proc Natl Acad Sci 102, 7988–7993 (2005). https://doi.org/10.1073/pnas.0409465102.

25) Li F. Receptor Recognition Mechanisms of Coronaviruses: a Decade of Structural Studies. J Virol 89, 1954–64 (2015). https://doi.org/10.1128/JVI.02615-14.

